# Inhaled combusted cannabis use is associated with proatherogenic changes in young people: A cross-sectional study

**DOI:** 10.64898/2026.03.04.26347657

**Authors:** Theodoros Kelesidis, Leila Fotooh Abadi, Lama Tamang Pratik, Katherine Hampilos, Reece Fong, Joshua Sanchez, Isabelle Ruedisueli, Jeffrey Gornbein, Ziva Cooper, Holly R Middlekauff

## Abstract

**Background:** Inhaled combusted cannabis and co-use of combusted cannabis and nicotine electronic cigarettes (nECIGs) are on the rise, yet their long-term cardiovascular risk is unclear due to the high prevalence of confounders in observational human studies. Using primary plasma and monocytes and a novel *ex vivo* mechanistic model of two early steps in atherogenesis, this study examined whether chronic combusted cannabis use is associated with atherogenic changes, as estimated by 1) monocyte transendothelial migration (MTEM), and 2) monocyte-derived foam cell formation (MDFCF), and whether nECIG co-use further amplifies this risk.

**Methods:** A cross-sectional parallel group comparison study was conducted in healthy adults (21-30 years) who chronically 1) used combusted cannabis, 2) co-used both combusted cannabis and nECIGs, and 3) were non-using controls. Using our *ex vivo* atherogenesis assay, primary outcomes of MTEM, MDFCF, and median fluorescence intensity (MFI) of the lipid-staining fluorochrome BODIPY were determined using primary plasma and autologous primary monocytes from participants. Using flow cytometry and the fluorochrome CELLROX, cellular oxidative stress (COS) in monocytes was determined.

**Results:** Of the 134 participants, 59 used cannabis, 26 co-used cannabis/nECIG, and 49 were non-using controls. The groups had similar age, sex, and race. Median MTEM was 1.13 fold greater in people who used cannabis compared to non-users 27.8% (IQR 26.1:29.2%) vs 24.5%, (IQR 22.9:27.4%), p<0.0001, and tended to be greater in people who co-used cannabis/nECIG by 1.22-fold 34.1%, (IQR 29.9:38.3%, p=0.17). Median MDFCF and MFI were also increased in people who used cannabis compared to non-users (MDFCF 36.3%, IQR 31.8:35.8%, vs 26.6%, IQR 23.8:25.8%, 1.36-fold and MFI 1163.8, IQR 1042.8:1155.0, vs 940.2 IQR 849.9:1101.4, 1.24-fold) and were further increased in people who co-used cannabis/nECIG (MDFCF 48.7%, IQR 37.3:52.4%, 1.34-fold, MFI 1433.7, IQR 1263.8:1686.4, 1.23-fold; all comparisons p<0.008). Foam cell formation, but not transendothelial migration, was strongly positively correlated with COS. All primary outcomes increased with greater frequency of cannabis and/or nECIG use.

**Conclusions:** In healthy young adults, exclusive cannabis use is associated with increased atherogenic properties of monocytes and plasma, and this atherogenic effect is further amplified by co-use of nECIGs.

---

In the United States, where cannabis (with >0.3% delta-9-tetrahydrocannabinol) is now legal for medical or non-medical use in thirty-nine states and the District of Columbia, 52.5 million adults (20% of the population) report past-year use (1). Legalization and the normalization of cannabis use have contributed to the perception that cannabis is safe (2). Contrary to this perception, and despite the fact that cannabis has been available and widely used for decades, it remains unclear whether cannabis increases long-term cardiovascular risk. Admittedly, there is an abundance of case reports in the medical literature describing an otherwise healthy young person with an acute myocardial infarction (MI) temporally linked with acute cannabis use (3, 4). Most large cohort studies, but not all, have reported increased MI risk in people with a cannabis use history (5–11). A recent meta-analysis, the first of its kind, reported an association between cannabis use and cardiovascular events, although the investigators highlighted several limitations of the studies included, particularly uncontrolled confounding factors (12). The greatest risk reportedly is within 60 minutes of use, consistent with the concept that cannabis may trigger an acute coronary event, even in the absence of known coronary artery disease (5). Similar to acute cocaine use, inhaled cannabis acutely increases sympathetic tone, and this sympathomimetic effect is one mechanism whereby cannabis (and cocaine) may trigger an acute ischemic event; other proposed mechanisms include coronary vasospasm, platelet activation and thrombosis, and/or microvascular dysfunction (3, 13–15).

However, as tragic as it is when an MI occurs in an otherwise healthy young person, this occurs relatively rarely (16). The greater threat to public health is the possibility that chronic use of inhaled cannabis leads to the development of coronary atherosclerosis. Yet even studies conducted in large data bases with the aim of determining long-term cardiovascular risk have led to inconsistent results, at least in part due to confounding factors, such as the fact that for many years, the people who smoked cannabis often also smoked tobacco cigarettes (7, 8, 10, 17, 18). Currently, due to public health campaigns, differential taxing, and laws prohibiting tobacco smoking in most public places, tobacco cigarette smoking has never been lower (19). In fact, the most popular tobacco product used by young adults in the United States is the nicotine electronic cigarette (nECIG) (19).

Atherosclerosis is a slowly progressive, inflammatory process that begins early in life with two early steps: (1) circulating monocyte transendothelial migration (MTEM) into the sub-endothelial space at sites of endothelial activation, and (2) monocyte phagocytosis of oxidized lipids leading to monocyte-derived foam cell formation (MDFCF) (20, 21). Foam cells, derived largely from monocytes but also from vascular smooth muscle cells and endothelial cells (22), accumulate in the sub-endothelial space, forming a fatty streak that is present in most Americans in their 20s (23). Foam cells secrete proinflammatory cytokines, ultimately leading to the progressive process of inflammatory atherosclerosis that, unchecked, may present decades later as an acute coronary syndrome (20). However, the direct proatherogenic role of cannabis use cannot be easily studied in observational human studies due to a lack of access to arterial tissue and the presence of several confounders that may also be proatherogenic. To address these knowledge gaps, we have established a mechanistic *ex vivo* model of atherogenesis the allows the presence of proatherogenic properties, including the MTEM and MDFCF, in human plasma and monocytes to be determined (24).

Using this novel *ex vivo* mechanistic atherogenesis assay, flow cytometry, and primary plasma and monocytes from otherwise healthy young people, including 1) people who used combusted cannabis, 2) people who co-used both combusted cannabis and nECIGs, and 3) people who were non-using controls, we tested the hypothesis that chronic combusted cannabis use increases two early steps in atherogenesis, MTEM and MDFCF, and that these changes are further increased in people who co-use combusted cannabis with nECIGs, the tobacco product of choice among young people.

## Methods

The data that support this study’s findings are available from the corresponding author upon reasonable request. The author has full access to the data and is responsible for data integrity and analysis.

### Study Population

As previously reported(25), this parallel group comparison study evaluated healthy male and female subjects aged of 21 to 30 years who do not smoke traditional cigarettes (TCIGs). Healthy male and female participants were eligible for enrollment in this parallel group comparison study if they: 1) were nonobese (≤30 kg/m2 body mass index); 2) had no known health problems (including asthma, diabetes, heart disease, hypertension, or hyperlipidemia); 3) were not competitive athletes; 4) had an alcohol intake ≤ 2 drinks per day; and 5) did not use illicit drugs other than cannabis, determined through self-report and urine toxicology testing. Based on their patterns of inhaled cannabis and nECIG use, participants were placed in one of the following three cohorts: 1) chronic (≥ one year, ≥ one time per week) inhaled combusted cannabis use, 2) chronic (≥ one year, ≥ one time per week) inhaled combusted cannabis and nECIGs use, or 3) non-using controls. Although a fourth group of people who exclusively used nECIGs was initially included, despite vigorous recruitment efforts, we enrolled only four individuals who exclusively used nECIGs and met other enrollment criteria. As a result, the exclusive nECIG group was excluded from the final analysis. Participants who used non-combusted forms of cannabis were eligible for the inhaled combusted cannabis group if they also regularly (≥ one time per week) used combusted cannabis. Former TCIG smoking was allowed if it had been at least one year since use. Participants were excluded from the study if they met the following exclusion criteria during the study visit: 1) non-sinus cardiac rhythm on ECG (e.g. atrial fibrillation); 2) positive urine pregnancy test; 3) positive urine toxicology test for any substance other than cannabis; or 4) end tidal carbon monoxide measurement > 10 ppm consistent with recent (within 8 hours) combusted cannabis use. The experimental protocol was approved by the Institutional Review Board at the University of California, Los Angeles (Los Angeles, California) and written informed consent was obtained from each participant. All procedures were followed in accordance with institutional guidelines.

### Phlebotomy

Blood samples were drawn by a trained medical assistant and sent to the UCLA Clinical Laboratory for measurement of plasma cotinine levels (half-life 36 hours). The commercial laboratory, ARUP Laboratories, performed this measurement using quantitative liquid chromatography–tandem mass spectrometry. Blood was also processed for isolation of plasma, cryopreserved at −80 °C, and peripheral blood mononuclear cells (PBMCs) cryopreserved at −196 °C.

### Ex Vivo Assay

Flow cytometry quantified two of the early steps in atherogenesis, MTEM and MDFCF, as previously described (24, 26, 27). An overview of the *ex vivo* atherogenesis assay is shown in Figure 1. Briefly, PBMCs were added to tumor necrosis factor–activated human umbilical vein endothelial cells monolayers on type I collagen gels to transmigrate for 1 hour in the presence of 10% v/v plasma in serum-free media and form foam cells over 48 hours as previously described (26, 27). Lipids present in the participant’s plasma drive *ex vivo* MDFCF. Flow cytometry was utilized to determine the number of CD45+ CD11b+ macrophages inside the collagen gel (a measure of MTEM) and the median fluorescence intensity (MFI) of BODIPY inside the transmigrated gel macrophages (a measure of lipid content in macrophages and MDFCF). MTEM is assessed as the number of CD11b+macrophages in collagen gel that have undergone TEM. MDFCF is assessed by the percentage of CD11b+macrophages in collagen gel that have become foam cells. The gating strategy has previously been described (24) and is shown in Figure 2.

**Figure 1.**
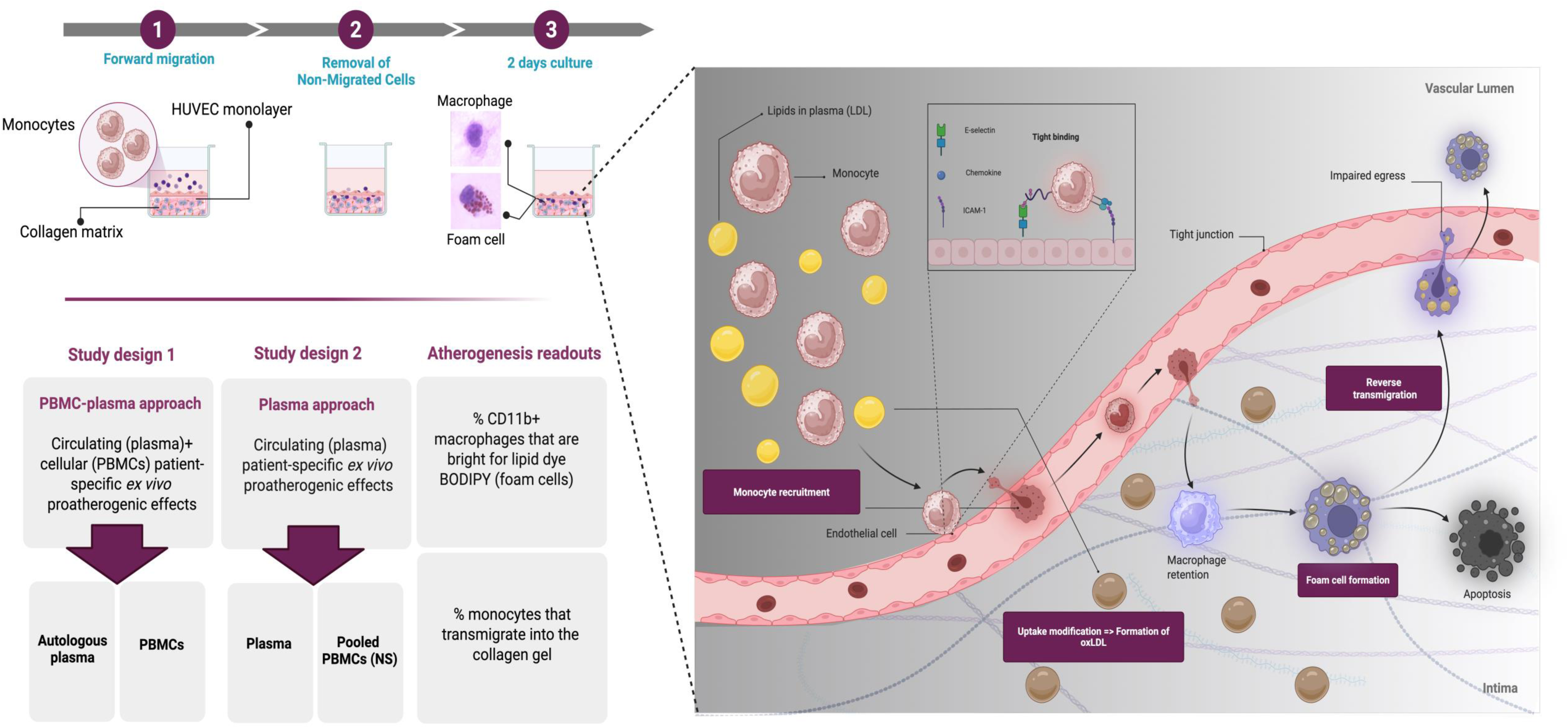
Study design and schema of the ex vivo atherogenesis modes.

**Figure 2:**
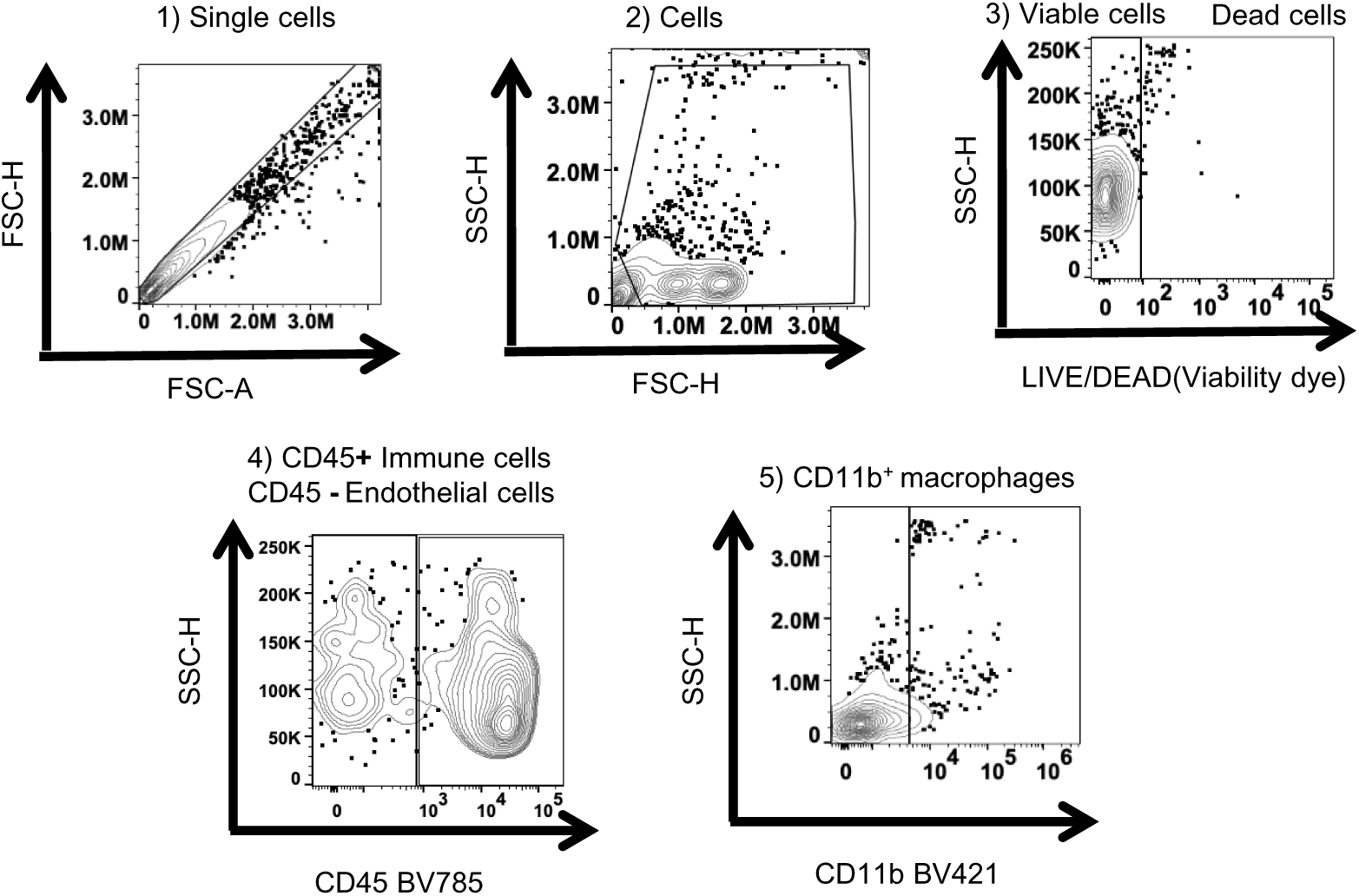
Flow cytometric strategy and gating in cells isolated from collagen gels in an ex vivo model of atherogenesis. The cells were gated sequentially as follows: **1) Gate 1 (singlets):** Single cells (x axis FSC-A, y axis FSC**); 2) Gate 2** (**cells; non-debris**) (x axis FSC-H, y axis SSC-H). Cells were gated from Gate 1 (singlets); **3) Gate 3 (viable cells):** x-axis Fluorescence viability dye-H, y-axis SSC-H. Viable cells were gated from Gate 2 (cells) as negative for the viability dye; **4) Gate 4 (cell subtypes):** x axis BV785-H (CD45), y axis SSC-H. Immune cells were gated from Gate 3 (viable cells) as CD45+ cells, and endothelial cells as CD45- cells. The collagen gels contain only immune and endothelial cells. **5) Gate 5 (macrophages):** x axis BV421-H (CD11b), y axis SSC-H. Macrophages were gated within the CD45+ gate (Gate 4) as CD45+/CD11b+.

To dissect circulating (present in plasma) and cellular (present in monocytes) participant-specific *ex vivo* proatherogenic effects, we utilized autologous participant plasma and PBMCs (“PBMC-plasma assay” in Figure 1). To dissect only circulating (present in plasma) participant-specific *ex vivo* proatherogenic effects, we utilized participant plasma and identical healthy non-smoking control PBMCs (“Plasma Assay” in Figure 1).

Additionally, in an exploratory analysis, we determined if endothelial cells also demonstrated increased lipid uptake when exposed to plasma and monocytes from people who used combusted cannabis or co-used cannabis/nECIGs. Accordingly, endothelial foam cell formation was assessed in our atherogenesis assay using two independent measures of foam cell formation in CD45- endothelial cells that had formed a monolayer on the collagen gel: 1) the % CD45- endothelial cells that had a bright positive signal for the lipid binding fluorochrome BODIPY (% of BODIPY bright+ of CD45- ECs), 2) the MFI of BODIPY in CD45- ECs.

To minimize experimental variability in an ex vivo model of atherogenesis, we employed a rigorous experimental approach: 1) Each biological replicate (from one unique human subject) was run in 4 technical replicates (collagen gels), 2) To minimize batch effects, an identical healthy non-smoking control PBMC with autologous plasma was included on each experimental plate, and all data were normalized to that control. The fold data were then transformed back to the original scale by multiplying by the average of that control group. 3) An identical number of samples per experimental group (cannabis, co-use, control) was included on each plate. Overall, the intra-assay coefficient of variation (CV%) and the inter-assay CV% were both < 15% for all experimental readouts.

### Cellular oxidative stress assay

Freshly isolated whole blood was immediately processed for flow cytometric determination of cellular ROS. Cellular oxidative stress (COS) was determined using the CellROX® Green Reagent, a measure of total cellular ROS (cytoplasmic and nuclear) as previously described (28).

### Urine Toxicology Screen

As previously reported (25, 29), at the beginning of the experimental session, participants provided a urine sample for immediate urine toxicology point-of-care testing for up to 12 drugs, including delta-9-tetrahydrocannabinol (THC) as well as amphetamines, benzodiazepines, cocaine, methamphetamines, opiates, and oxycodone (Alere iCup Dx Pro 2, Avatar).

### Questionnaire

The Cannabis Use Disorder Identification Test-Revised (CUDIT-R) (30), a validated screening tool for cannabis use disorder, was administered during the session. The CUDIT-R is useful to document patterns of cannabis consumption and its impact on social, psychological, and physical functioning.

### Study Session

Studies were conducted between 8 am and 2 pm. Participants were instructed to fast (except for water) and abstain from using nECIGs, cannabis, caffeine, or engaging in exercise for 8 hours before the study. Participants provided a urine sample at the beginning of the study and completed questionnaires describing their cannabis use patterns (30). Then, as previously described (31), participants were placed supine in a quiet, temperature-controlled (21°C) room in the Cannabis and Cannabinoid Research Laboratory in the UCLA Center for Cannabis and Cannabinoids. ECG electrodes were placed on the chest according to standard ECG protocol, and after a 10-minute rest period, blood pressure and heart rate were measured. Participants were not allowed to use their cell phones or talk during data acquisition. An ECG was then recorded for 5 minutes for later analysis of ventricular repolarization and heart rate variability. Results from the ECG and hemodynamic measurements have been previously reported (25). Blood was drawn, and the study was concluded. Investigators were not blinded to participant group during the experimental session but were blinded to group during performance of all analyses and assays of blood samples, which were performed later.

### Statistical Analysis

Using PBMCs and plasma from each participant (“PBMC-plasma assay,” Figure 1), the primary outcome measures were: 1) MTEM, % of blood monocytes that underwent transendothelial migration through a collagen gel, 2) MDFCF as determined by flow cytometry, and 3) the median fluorescence intensity (BODIPY MFI) of the lipid staining fluorochrome BODIPY in monocytes localized in the gel. In a secondary analysis to determine whether plasma contained atherogenic factor(s), these same outcomes were observed when the assay was performed with plasma from participants and PBMCs pooled from healthy non-using controls (“Plasma Assay,” Figure 1). Secondary outcomes included cellular oxidative stress (COS) measured in monocytes: 1) % CELLROX+ in CD14+ cells, and 2) MFI CELLROX in CD14+ cells.

We performed all analyses using R 4.3.2 (R Foundation for Statistical Computing, Vienna, Austria. <https://www.R-project.org/>). Fishers exact test was used to compute p values for comparing differences in sex and race/ethnicity across groups, and the nonparametric one-way Kruskal-Wallis procedure was used for comparing primary and most secondary continuous outcome measures across groups. The nonparametric Kruskal-Wallis was used since most outcomes did not follow a normal distribution, particularly in the tail area, although most distributions were roughly symmetric. The corresponding medians and interquartile ranges (IQR) are reported. Under the Kruskal-Wallis overall test, the Conover method was used to adjust p values to control for type I error among the three groups and an additional Bonferrori adjustment was made to control for the type I error rate among three primary outcomes (MTEM, MDFCF, MFI).

The association between two continuous variables was assessed using the nonparametric Spearman correlation (r_s_) since the Spearman correlation only assumes a monotone association, not a linear association. The Spearman correlation is reported for the association of a primary PBMC outcome (MTEM, MDFCF, MFI) versus CD 14 COS and CD14 COS MFI. Results are given stratified by group.

The simultaneous associations between a primary PBMC outcome versus CUDIT-R and cotinine adjusted for age, sex and positive vs negative urine THC were evaluated using multiple regression models for all groups combined. For MTEM and MDFCF, which are percentages between 0% and 100%, a multiple beta regression model was used which incorporates a logit (ln[Y/(100-Y)]) transformation for the outcome (Y) to avoid the “floor” effect of 0% and the “ceiling” effect of 100%. Since MFI followed a normal distribution on the log scale and was more linearly related to CUDIT-R and age on this scale, a multiple linear regression model was used with log_10_MFI as the outcome. Linearity was assessed using restricted cubic splines (RCS). Since cotinine was equal to zero in most controls and cannabis subjects (107 of 129 observations had cotinine=0) cotinine was collapsed into three ordered groups: 0, 1 to 100 and > 100 for the regression models.

For all models, the standardized regression coefficient and corresponding partial correlation is reported to quantify the association between the outcome versus the continuous cotinine or age. The standardized regression coefficient is the change in the outcome in standard deviation units for a one standard deviation increase in the cotinine, CUDIR-R or age predictor.

Differences or associations were considered statistically significant at a two-sided *p*<0.05 after Conover / Bonferroni adjustment. Regression results are exploratory and were not validated.

## Results

### Participant Baseline Characteristics

As previously reported(25), a total of 134 participants were enrolled in three groups, including 1) the control group (n=49), 2) the cannabis group (n=59), and 3) the cannabis/nECIG co-use group (n=26). As reported previously(25), groups did not differ significantly by sex (53% female), age (24.1± 2.9 years), or race/ethnicity (American Indian/Alaska Native 0.8%, Asian 34.3%, Black 5.2%, Hispanic 26.2%, White 23.1%, more than one or unknown 10.5%).

### Cannabis Use Characteristics

As previously reported (25), there were 85 participants who regularly used combusted cannabis with or without nECIGs, and smoking cannabis was the primary method in the majority (n=53; 62.4%) followed by vaping (n=12; 14.1%). Most used cannabis four or more times per week (n=42; 49.4%), followed by two to three times per week (n=26; 30.6%). Cannabis use burden was similar between exclusive cannabis and cannabis/nECIG co-use groups as estimated by their mean CUDIT-R score (12.7 ± 5.6 vs. 13.1 ± 6.3, *p*=0.939) and urine positivity for Δ9-tetrahydrocannabinol (THC, 61.5% vs. 49.2%, *p*=0.337). The median blood cotinine level in the cannabis/nECIG co-use group was 65.5 ng/ml (IQR 10.3-140.0 ng/ml), indicative of regular nECIG use, and was 0 ng/ml (interquartile range [IQR] 0-0 ng/ml) in the other two groups.

### Atherogenesis Assay: PBMCs and Plasma

In this set of experiments, plasma and PBMCs from each participant were used (“PBMC-plasma assay” in Figure 1) to determine whether chronic combusted cannabis use increases atherosclerosis risk, as estimated by 1) monocyte transendothelial migration (MTEM), and 2) monocyte-derived foam cell formation (MDFCF), and whether nECIG co-use further amplifies this risk.

*Proatherogenic MTEM.* First, we measured MTEM in our *ex vivo* model of atherogenesis. Representative data from high- and low-MTEM samples are shown in Figure 3A. The median MTEM was significantly greater in people who exclusively used cannabis by 1.13-fold compared to non-using healthy controls (27.8% IQR 26.1:29.2% vs 24.5%, IQR 22.9:27.4%, p<0.0001) (Figure 3B). The median MTEM tended to be even greater in the people who co-used cannabis/nECIG by of 1.22 fold compared to people who only used cannabis (34.1%, IQR 29.9: 38.3%, p=0.17) and had a median increase of 1.39-fold compared to non-using healthy controls (p<0.0004) (Figure 3B).

**Figure 3:**
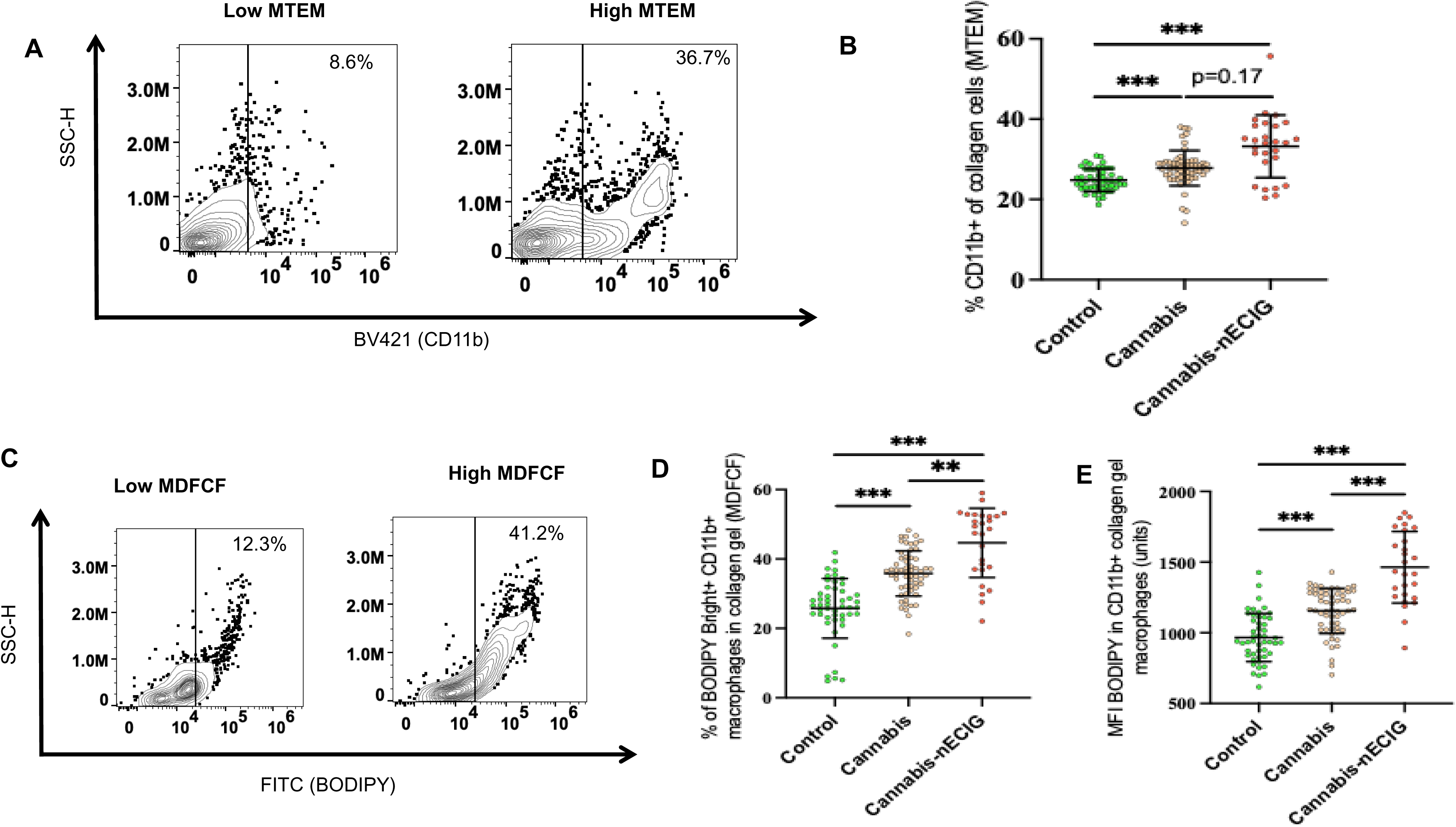
Differences in outcomes of ex vivo atherogenesis with the PBMC-plasma approach among groups with different substance exposure. Autologous plasma and autologous cells [peripheral blood mononuclear cells (PBMCs)] from each study participant from 3 groups [non-using controls (Control), n= 57, people who chronically exclusively use cannabis (Cannabis), n= 59, and people who chronically co-use nicotine ECIG and cannabis (Cannabis/nECIG), n= 27] were utilized in an ex vivo model of atherogenesis over 48 hours as described in methods. Monocyte transendothelial migration (MTEM) and monocyte-derived foam cell formation (MDFCF) were determined by flow cytometry in the setting of exposure of participant peripheral blood mononuclear cells (PBMCs) to autologous primary plasma as described in the methods. MTEM was determined by flow cytometry by measuring the % of CD11b+ macrophages in the collagen gel overlayed by a monolayer of endothelial cells **(A, B).** MDFCF was determined by flow cytometry by determination of two measures: **i)** the % of CD11b+ macrophages that were present in the collagen gel that had avidly taken up plasma lipids to become BODIPY Bright foam cells **(C, D),** ii) **i)** the median fluorescence intensity (MFI) of the lipid binding fluorochrome BODIPY in CD11b+ macrophages that were present in the collagen gel **(E)**, Representative data from MTEM (A) and MDFCF (C) are shown. Summary data from MTEM (C) and MDFCF (D, E) are also shown. The mean ± standard deviation (SD) for each measure is shown. The p-values for comparisons among the three groups were computed using the non-parametric Kruskal-Wallis method. The p-values were adjusted using the Tukey criteria for 3 groups. ***p<0.001 indicates statistically significant group differences.

*Proatherogenic MDFCF.* Second, we measured MDFCF in our *ex vivo* model of atherogenesis using two independent measures of foam cell formation in CD11b+ macrophages that have migrated through MTEM inside the collagen gel: i) the % CD11b+ macrophages that had a bright positive signal for the lipid binding fluorochrome BODIPY (% of BODIPY bright+ of CD11b+ monocytes), ii) the MFI of BODIPY in CD11b+ macrophages. Representative data from high- and low-MDFCF samples are shown in Figure 3C. The median % of BODIPY bright+ of CD11b+ cells was significantly greater in people who exclusively used cannabis by 1.36-fold compared to non-using healthy controls MDFCF (36.3% IQR 31.8:35.8% vs 26.6, IQR 23.8:30.7%, p<0.00001) (Figure 3D). The median % of BODIPY bright+ of CD11b+ cells was even greater in people who co-used cannabis/nECIGs by 1.34-fold compared to people who only used cannabis (48.7%, IQR 37.3:52.4 p<0.008), and by 1.83-fold compared to non-using healthy controls (p<0.00001) (Figure 3D). The median BODIPY MFI in macrophages was greater in people who exclusively used cannabis by 1.24-fold compared to non-using healthy controls (1163.8, IQR 1042.8:1286.0 vs 940.2 IQR 849.9:1101.4, p<0.00001) (Figure 3E). The median BODIPY MFI in macrophages was even greater by 1.23-fold in people who co-used cannabis/nECIGs compared to people who only used cannabis (1433.7, IQR 1263.8:1686.4, p<0.00001) and by 1.52-fold compared to non-using healthy controls(p<0.00001) (Figure 3E).

### Isolating the atherogenic contribution from plasma: Using participant specific plasma and control PBMCs

To dissect only circulating (present in plasma) participant-specific *ex vivo* proatherogenic effects, we then used participant plasma and identical healthy non-smoking control PBMCs (“Plasma Assay” in Figure 1).

*Proatherogenic MTEM.* When the assay was performed using plasma from participants but PBMCs pooled from healthy controls, the MTEM was not elevated in individuals who exclusively used cannabis compared to non-using healthy controls (21.9%, IQR 17.8:23.9% vs 20.6%, IQR 17.7:23.1%, p=0.73) (Figure 4A). The median MTEM tended to be higher in individuals who co-used cannabis/nECIG, with a 1.09-fold increase compared to those who only used cannabis (23.8%, IQR 22.0:26.9%, p=0.10, Figure 4A), and was higher by 1.16-fold, compared to non-using healthy controls (p=0.0026).

**Figure 4:**
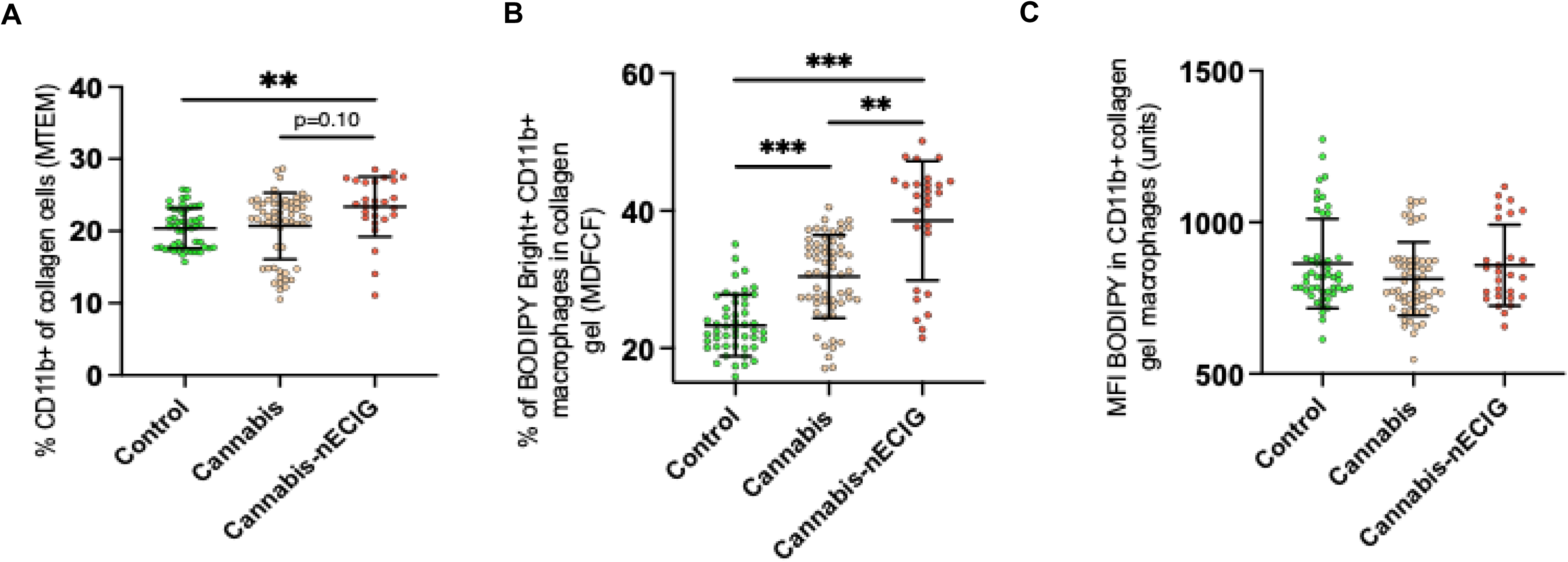
Differences in outcomes of ex vivo atherogenesis with plasma approach among groups with different substance exposure. Primary plasma from each study participant from 3 groups [non-using controls (Control), n= 57, people who chronically exclusively use cannabis (Cannabis), n= 59, and people who chronically co-use nicotine ECIG and cannabis (Cannabis/nECIG), n= 27] and healthy cells [pooled peripheral blood mononuclear cells (PBMCs) from control group] were utilized in an ex vivo model of atherogenesis over 48 hours as described in methods (Approach B). Monocyte transendothelial migration (MTEM) and monocyte-derived foam cell formation (MDFCF) were determined by flow cytometry in the setting of healthy PBMCs exposed to primary plasma, as described in the methods. MTEM was determined by flow cytometry, measuring the % of CD11b+ macrophages in a collagen gel overlayed with a monolayer of endothelial cells **(A)**. MDFCF was determined by flow cytometry by determination of two measures: **i)** the % of CD11b+ macrophages that were present in the collagen gel that had avidly taken up plasma lipids to become BODIPY Bright foam cells **(B)**, ii) the median fluorescence intensity (MFI) of the lipid binding fluorochrome BODIPY in CD11b+ macrophages that were present in the collagen gel **(C)**. The mean ± standard deviation (SD) for each measure is shown. The p-values for comparisons among the three groups were computed using the non-parametric Kruskal-Wallis method. The p-values were adjusted using the Tukey criteria for 3 groups. *p<0.05, **p<0.01, ***p<0.001, ****p<0.0001 indicate statistically significant group differences.

*Proatherogenic MDFCF.* When the assay was performed using plasma from participants but PBMCs pooled from healthy controls, median MDFCF was greater in people who exclusively used cannabis by 1.38-fold compared to non-using healthy controls (31.2%, IQR 26.9:35.0% vs 22.6%, IQR 20.3:26.4%, p< 0.00001) (Figure 4B). Median MDFCF was even greater in people who co-used cannabis/nECIG by 1.35-fold compared to people who only used cannabis (42.2%, IQR 32.6:44.1%, p=0.0051), and by 1.87-fold compared to non-using healthy controls (p<0.00001) (Figure 4B).

*Proatherogenic BODIPY-MFI.* When the assay was performed using plasma from participants but PBMCs pooled from healthy controls, BODIPY-MFI was not greater in people who exclusively used cannabis (781.3, IQR 731.8:873.2) or in people who co-used cannabis/nECIG (826.4, IQR 754.9:949.2) compared to healthy non-using controls (818.4, IQR 774.7:883.1; overall p value 0.14) (Figure 4C).

Overall, changes in *ex vivo* atherogenesis readouts (MTEM, MDFCF) were greater in the PBMC-plasma approach than in the plasma-only approach in the cannabis-use groups compared to the control group, suggesting that cannabis use had both cellular and circulating proatherogenic effects on monocytes rather than just systemic/circulating effects.

### Endothelial cell foam cell formation

Endothelial cells have been identified as a source of foam cell formation in atherogenic plaques, although studies on this are limited (22, 32–34). Therefore, in an exploratory analysis to determine if endothelial cells also showed increased lipid uptake when exposed to plasma and monocytes from people who used combusted cannabis or co-used cannabis/nECIGs, we assessed foam cell formation in our atherogenesis assay using two independent measures in CD45- endothelial cells that formed a monolayer on collagen gel: i) the percentage of CD45-endothelial cells (ECs) that exhibited a bright positive signal for the lipid-binding fluorochrome BODIPY (percentage of BODIPY bright+ of CD45- ECs), and ii) the mean fluorescence intensity (MFI) of BODIPY in CD45- ECs.

When the assay was performed with the PBMC-Plasma assay, the median % of BODIPY bright+ of CD45- ECs was not different in people who exclusively used cannabis compared to non-using healthy controls (12.2%, IQR 8.8:12.9% vs 9.5%, IQR 7.4:12.8 p=0.20) (Figure 5A). The median % of BODIPY bright+ of CD45- ECs was higher in people who co-used cannabis/nECIGs by 1.12-fold compared to people who only used cannabis (13.7%, IQR 12.0: 23.3%, p=0.013) and by 1.46-fold compared to non-using healthy controls (p<0.00001) (Figure 5A). The BODIPY MFI in CD45- ECs was not different in people who exclusively used cannabis compared to non-using healthy controls (309.0, IQR 288.8:322.4 vs 289.6 IQR 260.5:311.2, p=0.11) (Figure 5B). The median BODIPY MFI in CD45- ECs was higher by 1.25-fold in people who co-used cannabis/nECIGs compared to people who only used cannabis (386.8, IQR 378.0:436.7, p<0.00001), and by 1.34-fold compared to non-using healthy controls (p<0.00001) (Figure 5B).

**Figure 5:**
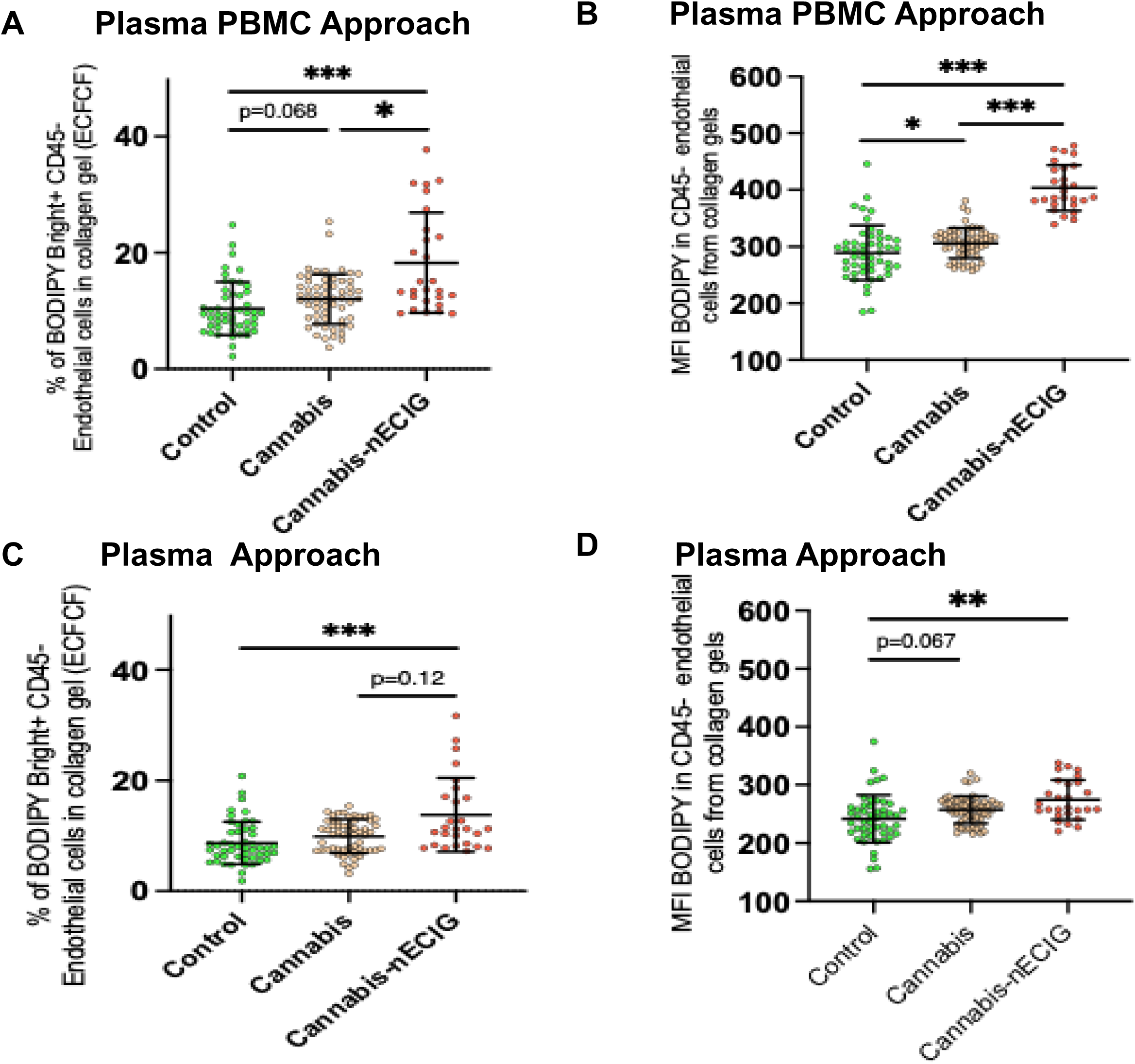
Differences in endothelial cell-derived foam cell formation (ECFCF) among groups with different substance exposure. **A, B:** Autologous plasma and autologous cells [peripheral blood mononuclear cells (PBMCs)] from each study participant from 3 groups [non-using controls (Control), n= 57, people who chronically exclusively use cannabis (Cannabis), n= 59, and people who chronically co-use nicotine ECIG and cannabis (Cannabis/nECIG), n= 27] were utilized in an ex vivo model of atherogenesis over 48 hours as described in methods. Endothelial cell-derived foam cell formation (ECFCF) was determined by flow cytometry in the setting of participant peripheral blood mononuclear cells (PBMCs) exposure to autologous primary plasma as described in the methods. ECFCF was determined by flow cytometry by determination of two measures: **i)** the % of CD45- endothelial cells that were present in the collagen gel that had avidly taken up plasma lipids to become BODIPY Bright foam cells **(B)**, ii) **i)** the median fluorescence intensity (MFI) of the lipid binding fluorochrome BODIPY in CD45-endothelial cells that were present in the collagen gel. **C, D:** Primary plasma from each study participant in 3 groups and healthy cells (PBMCs from the control group) were used in an ex vivo model of atherogenesis over 48 hours, as described in the methods (Plasma Assay). ECFCF was detected in **A and B.** The mean ± standard deviation (SD) for each measure is shown. The p-values for comparisons among the three groups were computed using the non-parametric Kruskal-Wallis method. The p-values were adjusted using the Tukey criteria for 3 groups. *p<0.05, **p<0.01, ***p<0.001, ****p<0.0001 indicate statistically significant group differences.

As we did with the monocytes, we then used participant plasma and pooled healthy non-smoking control PBMCs to dissect only circulating (present in plasma) participant-specific *ex vivo* proatherogenic effects (“Plasma Assay” in Figure 1).

When the assay was performed with the plasma only method, the % of BODIPY bright+ of CD45- ECs was not greater in people who exclusively used cannabis compared to non-using healthy controls (10.5%, IQR 7.4:12.7 vs 7.8, IQR 6.2:10.8%, p= 0.23) (Figure 5C). The median % of BODIPY bright+ of CD45- ECs tended to be higher in people who co-used cannabis/nECIG by 1.08-fold compared to people who only used cannabis (11.3%, IQR 8.4:17.0%, p=0.12), and by 1.4-fold compared to non-using healthy controls (p=0.0004) (Figure 5C).

The BODIPY MFI in CD45- ECs tended to be greater in people who exclusively used cannabis by a mean of 1.07-fold compared to non-using healthy controls (259.7, IQR 242.6:271.0 vs 243.3, IQR 217.2:261.4, p=0.067) (Figure 5D). The median BODIPY MFI in CD45- ECs was not greater in people who co-used cannabis/nECIG compared to people who only used cannabis (260.9, IQR 253.4:302.8, p=0.55) but was greater by 1.2-fold compared to non-using healthy controls (p=0.0024) (Figure 5D).

Overall, as with monocytes, all changes in endothelial cells in *ex vivo* atherogenesis measures were greater in the PBMC-plasma approach than in the plasma-only approach in the cannabis-use groups compared to the control group, suggesting that cannabis use had both cellular and circulating, not just circulating, proatherogenic effects on endothelial cells.

### Cellular Oxidative Stress (COS)

Reactive oxygen species (ROS) and cellular oxidative stress (COS) in CD14+ monocytes are established instigators of atherogenesis(35). To determine whether cannabis use contributes to atherogenesis by altering COS in CD14+ monocytes, we measured COS by flow cytometry using the fluorochrome CELLROX, in association with our *ex vivo* measures of atherogenesis. We determined COS using two independent measures: i) the % CELLROX+ of cells, ii) the CELLROX MFI in cells.

COS measured by % CELLROX+ of CD14^+^ cells was significantly higher in people who exclusively used cannabis, with a median increase of 1.50 times compared to non-using healthy controls (42.1%, IQR 39.1:44.7% vs 28.1%, IQR 25.6:31.5%, p<0.00001) (Figure 6A). The % CELLROX+ of CD14^+^ cells was also higher in people who co-used cannabis/nECIG, with a median increase of 1.38 times compared to those who only used cannabis (57.9%, IQR 51.9:60.7%, p<0.00001) and a 2.06-fold increase compared to non-using healthy controls (p<0.00001, Figure 6A).

**Figure 6:**
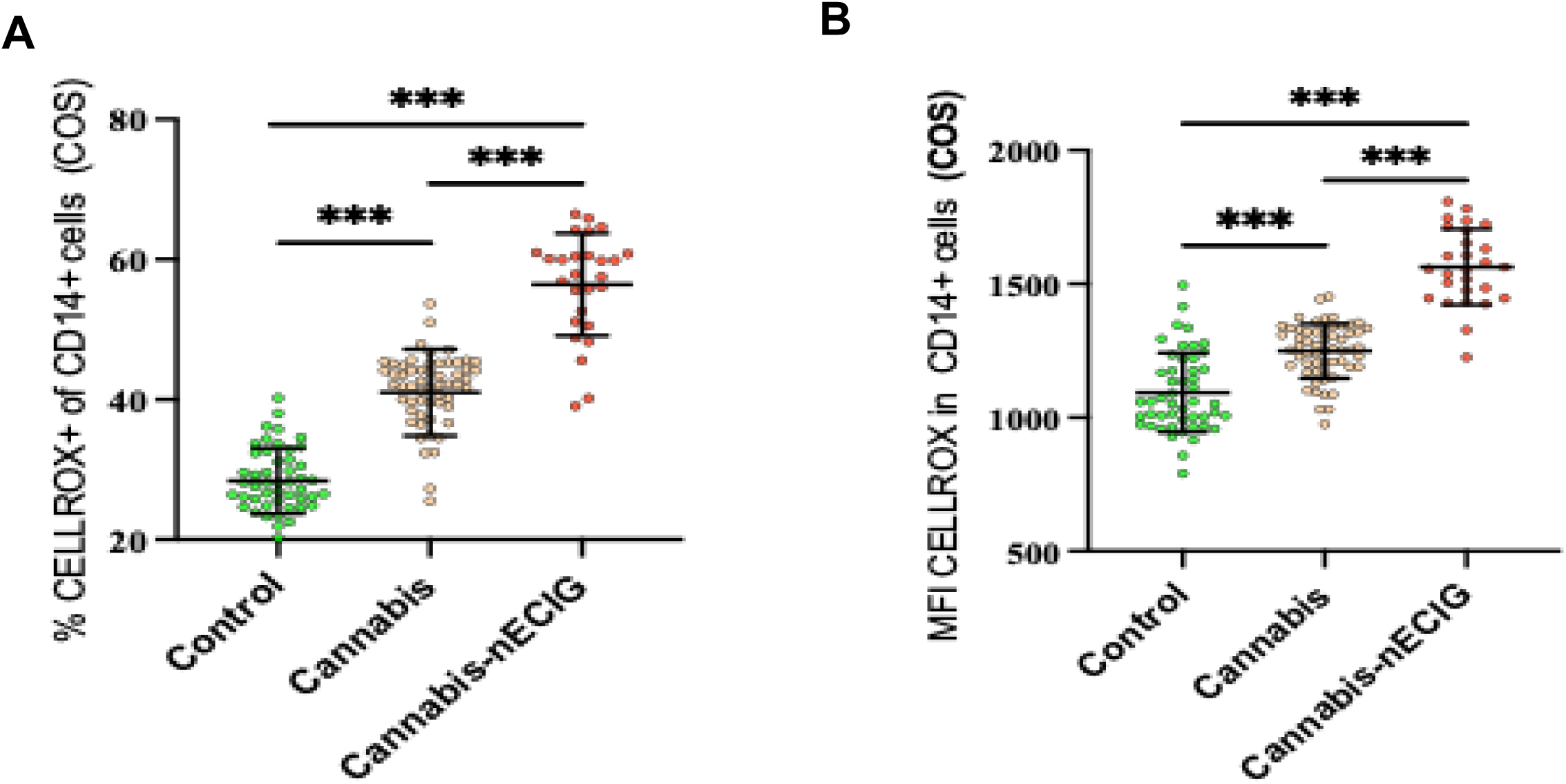
Differences in measures of cellular oxidative stress (COS) in immune cells isolated from groups with different substance exposure. Peripheral blood mononuclear cells (PBMCs) from each study participant from 3 groups [non-using controls (Control), n= 57, people who chronically exclusively use cannabis (Cannabis), n= 59, and people who chronically co-use nicotine ECIG and cannabis (Cannabis/nECIG), n= 27] were utilized to determine cellular oxidative stress by flow cytometry as described in the methods. COS was determined by flow cytometry in CD14+ monocytes **(A, B)** by determination of two measures: **i)** the % of cells that were positive for the fluorochrome CELLROX (that detects reactive oxygen species), **ii)** the median fluorescence intensity (MFI) of the fluorochrome CELLROX in cells. The mean ± standard deviation (SD) for each measure is shown. The p values for comparisons among the three groups were computed using the non-parametric Kruskal-Wallis method. The p values were adjusted using the Tukey criteria for 3 groups. ***p<0.001, ****p<0.0001 indicate statistically significant group differences.

COS measured as MFI in CD14+ cells was significantly greater in people who exclusively used cannabis by a median of 1.19-fold compared to non-using healthy controls (1255.1, IQR 1191.6:1325.9 vs 1059.7, IQR 992.5:1180.9, p<0.00001) (Figure 6B). MFI in CD14+ cells was even greater in people who co-used cannabis/nECIG by a median of 1.24-fold compared to people who only used cannabis (1555.1, IQR 1461.3:1676.8, p<0.00001) and by a median of 1.47-fold compared to non-using healthy controls (p<0.00001) (Figure 6B).

In summary, using two different independent measures of COS, the % CELLROX+ of CD14+ cells and the CELLROX MFI in CD14+ cells, COS was greater in people who used combusted cannabis and this effect was amplified by nECIG co-use.

### Associations between measures of cellular oxidative stress and primary outcomes of atherogenesis

To further explore our hypothesis that proatherogenic effects of cannabis are mediated through systemic effects on oxidative stress, we then determined the associations of these two measures of COS, % CELLROX+ of CD14^+^ cells, and MFI in CD14+ cells, with the primary and secondary outcomes of atherogenesis. For this exploratory analysis, we focused on the outcomes of *ex vivo* atherogenesis using only the PBMC-plasma approach, which includes cells and plasma from each participant (so we can link the COS from each participant to the *ex vivo* atherogenesis readouts from the same participant). The association between COS measures and *ex vivo* atherogenesis is shown in Table 1.

**Table 1.** Spearman correlations between primary outcomes from ex-vivo atherogenesis assay and cellular oxidative stress assay in immune cells by group.

| **Control** | **MTEM** |  | **MDFCF** |  | **BODIPY MFI** |  |
| --- | --- | --- | --- | --- | --- | --- |
|  | **Spearman** |  | **Spearman** | **p-** | **Spearman** | **p-** |
|  | **correlation** | **p-value** | **correlation** | **value** | **correlation** | **value** |
| **CD14+ COS** | -0.722 | <0.0001 | 0.791 | <0.0001 | 0.812 | <0.0001 |
| **CD14+ COS MFI** | -0.732 | <0.0001 | 0.791 | <0.0001 | 0.804 | <0.0001 |
| **Exclusive Cannabis** | **MTEM** |  | **MDFCF** |  | **BODIPY MFI** |  |
|  | **Spearman** |  | **Spearman** | **p-** | **Spearman** | **p-** |
|  | **correlation** | **p-value** | **correlation** | **value** | **correlation** | **value** |
| **CD14+ COS** | -0.378 | 0.0035 | 0.208 | 0.12 | 0.501 | 0.0001 |
| **CD14+ COS MFI** | -0.460 | 0.0003 | 0.357 | 0.006 | 0.756 | <0.0001 |
| **Cannabis/nECIG** | **MTEM** |  | **MDFCF** |  | **BODIPY MFI** |  |
|  | **Spearman** |  | **Spearman** | **p-** | **Spearman** | **p-** |
|  | **correlation** | **p-value** | **correlation** | **value** | **correlation** | **value** |
| **CD14+ COS** | 0.052 | 0.797 | 0.220 | 0.268 | 0.432 | 0.026 |
| **CD14+ COS MFI** | 0.081 | 0.686 | 0.184 | 0.357 | 0.648 | 0.0004 |

In the control group, both measures of COS had a strong negative association with MTEM, (% CELLROX+ of CD14^+^ immune cells: r_s_= -0.722, p<0.0001; MFI of CD14+: r_s_=-0.732, <0.0001), and a strong positive association with MDFCF (% CELLROX+ of CD14^+^ immune cells: r_s_=0.791, p<0.0001; MFI of CD14+: r_s_=0.791, p<0.0001), and BODIPY MFI (% CELLROX+ of CD14^+^ immune cells: r_s_=0.812, p<0.0001; MFI of CD14+: 0.804, p<0.0001, (Table 1).

In the exclusive cannabis group, both measures of COS had a weak negative association with MTEM, (% CELLROX+ of CD14^+^ immune cells: r_s_=-0.378, p=0.0035; MFI of CD14+: r_s_=-0.460, p=0.0003), a weak positive association with MDFCF (% CELLROX+ of CD14^+^ immune cells: r_s_=0.208, p=0.12; MFI of CD14+: 0.357, p=006), and a strong positive association with BODIPY MFI (% CELLROX+ of CD14^+^ immune cells: r_s_=0.501, p=0.0001; MFI of CD14+: r_s_=0.756, p<0.0001), (Table 1)

In the cannabis/nECIG co-use group, COS levels were not significantly associated with MTEM, (% CELLROX+ of CD14^+^ immune cells: r_s_=0.052, p=0.791; MFI of CD14+: r_s_=0.081, p=0.686),

or MDFCF, (% CELLROX+ of CD14^+^ immune cells: r_s_=0.220, p=0.268; MFI of CD14+: r_s_=0.184, p=0.357), but had a moderate to strong positive association with BODIPY MFI, (% CELLROX+ of CD14^+^ immune cells: r_s_=0.432, p=0.026; MFI of CD14+: r_s_=0.648, p=0.0004), (Table 1)

In summary, greater levels of COS were most strongly associated with greater foam cell formation, a process in which lipid oxidation is critical (35).

### Multiple Regression of Primary Outcomes (MTEM, MDFCF, BODIPY-MFI)

Our multiple regression model included continuous predictors of age, CUDIT-R score (an estimate of cannabis use and risk for cannabis use disorder), and plasma cotinine level (an estimate of nECIG use), and the binary predictors of urine toxicology testing (positive or negative for cannabis), and self-reported biological sex. Only CUDIT-R scores and plasma cotinine levels were independently associated with atherogenesis. Specifically, greater use of combusted cannabis, as estimated by the CUDIT-R score, was associated with greater MDFCF and MFI (Table 2). While there was also a positive association between CUDIT-R and MTEM, it did not reach statistical significance (partial r=0.129, p=0.1545). Further, greater use of nECIGs, as estimated by plasma cotinine level, was also associated with greater MTEM, MDFCF and MFI (Table 2). These findings support the concept that combusted cannabis increases atherogenic risk, and that co-use of nECGs amplifies this risk.

**Table 2.** Multivariable regression analysis of primary PBMC outcomes from ex-vivo atherogenesis assay of participant peripheral blood mononuclear cells and plasma.

|  | MTEM |  |  | MDFCF |  |  | BODIPY MFI |  |  |
| --- | --- | --- | --- | --- | --- | --- | --- | --- | --- |
|  | Std rate | Partial | p value | Std rate | Partial corr | p value | Std rate | Partial | p |
| **Continuous predictor** |  |  |  |  |  |  |  |  |  |
| CUDIT-R | 0.150 | 0.129 | 0.1545 | 0.430 | 0.376 | < 0.001 | 0.385 | 0.367 | < 0.001 |
| Age (per yr) | -0.154 | -0.160 | 0.0753 | 0.082 | 0.094 | 0.2983 | 0.080 | 0.099 | 0.2735 |
|  | **median** | **p value** |  | **median** | **p value** |  | **median** | **p value** |  |
| **Blood Cotinine level (ng/ml)** |  |  |  |  |  |  |  |  |  |
| 0 | 27.1 | reference |  | 30.8 | reference |  | 1063.3 | reference |  |
| 1-100 | 30.7 | 0.0447 |  | 39.7 | 0.0249 |  | 1257.7 | 0.0065 |  |
| 101+ | 30.8 | 0.0242 |  | 43.5 | 0.0007 |  | 1507.0 | < 0.0001 |  |
|  | **Median difference** | **p value** |  | **Median difference** | **p value** |  | **Median difference** | **p value** |  |
| **Urine (pos vs neg)** | 1.14% | 0.3704 |  | -0.69% | 0.8093 |  | -22.6 | 0.6729 |  |
| **Male vs female** | 0.17% | 0.8615 |  | -0.02% | 0.9909 |  | -6.7 | 0.8684 |  |
| **overall corr \*** | 0.407 |  |  | 0.603 |  |  | 0.644 |  |  |

\* overall Spearman correlation between model predicted vs observed PBMC outcome
Values are model-adjusted standardized rates (standardized regression coefficients) and corresponding partial correlations (partial corr) for continuous predictors, and model-adjusted medians or median differences for categorical predictors. Participant group (i.e., control, exclusive cannabis, ECIG/cannabis co-use) was not included as a potential predictor, as the combination of THC urine test, blood cotinine, and CUDIT-R score defines groups.

## Discussion

Cannabis is a complex drug with opposing effects on the immune system, primarily mediated by different cannabinoid (CB) receptors (36). Preclinical studies have shown that CB-1 receptors, which are widely distributed throughout the body—including on cardiomyocytes, central and peripheral neurons, immune cells, endothelial cells, and others—have pro-oxidative and pro-inflammatory effects. In contrast, CB-2 receptors, found on immune cells, reduce the formation of reactive oxygen species and have anti-inflammatory effects (13, 37–39). Therefore, in people who regularly use combusted cannabis, both the anti-oxidative, anti-inflammatory pathways and the pro-oxidative, pro-inflammatory pathways are active. The overall net effect, and its implications for the development of future inflammatory atherosclerosis, remain uncertain (17). However, as cannabis use becomes more common, understanding whether chronic use of combusted cannabis is associated with long-term proatherogenic effects is increasingly urgent. As a result, this study’s findings offer much-needed clarity.

Using a unique translational approach with a mechanistic *ex vivo* model of atherogenesis, we provide, to our knowledge, among the first mechanistic evidence in humans that chronic combusted cannabis use is associated with proatherogenic effects that may contribute to long-term cardiovascular risk. We demonstrated that: 1) as measured in our *ex vivo* atherogenesis assay, transendothelial monocyte migration and foam cell formation, two early steps in inflammatory atherosclerosis, are increased in people who chronically use combusted cannabis; 2) these early steps in atherogenesis are further increased in people who co-use nicotine electronic cigarettes with combusted cannabis compared to those who only use combusted cannabis; 3) overall, atherogenic effects (MTEM, MDFCF) were higher in the PBMC-plasma approach compared to the plasma-only approach, suggesting cannabis use impacts both cellular and circulating proatherogenic effects on monocytes; 4) increased cellular oxidative stress is strongly linked with foam cell formation but not transendothelial migration of monocytes in people who smoke cannabis; 5) in this healthy young cohort, greater cannabis use frequency correlates with greater pro-atherogenic effects in our assay, and these effects are independently and further heightened with increased frequency of nECIG use, indicating a harm amplification effect when co-using nECIG; and finally, in an exploratory analysis, 6) combusted cannabis use raises lipid uptake in endothelial cells (EC), contributing to pro-atherogenic effects, and this EC-derived foam cell formation is even greater in people who co-use cannabis and nECIG, suggesting diverse cell types contribute to cannabis-related pro-atherogenic effects; this observation warrants further investigation (22, 32–34). In summary, this study adds to the growing evidence that frequent cannabis smoking is associated with pro-atherogenic changes that may lead to inflammatory atherosclerosis.

Mohammadi and colleagues recently reported that healthy, non-smoking adults under age 50 who chronically use combusted or oral THC-dominant cannabis exhibit significant endothelial dysfunction as estimated by brachial-artery flow-mediated dilation (FMD) compared to non-cannabis users (40). Abnormal FMD, driven by endothelial dysfunction, is one of the earliest signs of future atherosclerosis and has been linked to the development of adverse cardiovascular events (41, 42). Increased oxidative stress has been identified as a key mechanism underlying reduced FMD (43, 44). Our findings show that monocytes from otherwise healthy young people who regularly use combusted cannabis have increased atherogenic properties, including higher migration into the subendothelial space and greater foam cell formation, which correlate with increased oxidative stress, and further support this phenotypic evidence of cannabis-induced vascular dysfunction. Specifically, our results provide mechanistic evidence that chronic combusted cannabis use is associated with proatherogenic changes that may lead to inflammatory atherosclerosis. Interestingly, in an exploratory analysis, we also assessed the potential for endothelial cells to produce foam cells in our model, where healthy endothelial cells were exposed to plasma and monocytes from people who use combusted cannabis and co-use cannabis/nECIGs. Consistent with Mohammadi and colleagues’ findings that chronic cannabis use is associated with proatherogenic endothelial dysfunction (40), we demonstrated that cannabis use is linked to proatherogenic lipid uptake by endothelial cells in an ex vivo setting, and this effect is amplified with co-use of nECIGs. Indeed, research on EC-derived foam cells remains limited, and the mechanisms of abnormal lipid metabolism involved in foam cell formation require further investigation (22).

For many years, a large proportion of people who smoked cannabis also smoked tobacco cigarettes, but currently, combusted tobacco use is at historic lows, while nECIGs have gained popularity. The prevalence of co-use of cannabis and nECIGs has been reported to exceed 17% in young people(45). Our findings demonstrate that adding chronic nECIG use to combusted cannabis use significantly increases atherogenic potential compared to only using combusted cannabis. This finding is consistent with our previous work, in which we reported that atherogenic properties of monocytes and plasma were increased in people who chronically and exclusively used nECIGs, compared to non-smoking controls(24). Interestingly and perhaps not surprisingly, in that study, people who chronically and exclusively smoked tobacco cigarettes had even higher atherogenic potential than people who exclusively used nECIGs; people who were dual users (tobacco cigarettes and nECIGs) were not studied. The remarkable finding in the current study is that the effect of nECIGs is not obscured by the combusted cannabis use, consistent with an additive effect, perhaps via activation of distinct inflammatory or atherogenic pathways.

Repeating the assay with monocytes from non-using controls and plasma from individuals who exclusively use cannabis or co-use with nECIGs is insightful. When the assay was performed using plasma from people who used cannabis or co-used cannabis/nECIGs, the addition of autologous monocytes from those groups consistently increased atherogenesis as measured by MTEM and MDFCF. Additional studies to identify the cellular pathways activated in monocytes from these groups compared to non-users, in order to uncover differences, may be revealing and are needed.

In our multivariate analysis, increased monocyte transendothelial migration and foam cell formation were significantly associated with increased use of combusted cannabis and/or nECIGs. These findings are consistent with our hypothesis that combusted cannabis is associated with pro-atherogenic properties, which are amplified by nECIG co-use. Interestingly, increasing age did not confer increasing risk. This is likely attributable to our study population’s very narrow, young age range and to the fact that we are studying early markers, the purported *instigators* of inflammatory atherosclerosis. If an outcome were studied that was associated with more advanced atherosclerosis, for example, coronary calcification as detected by CT scan, in a population with a wider age range, then age may also have been an independent risk factor. Although sex did not influence the proatherogenic associations with combusted cannabis use, this apparent similarity between the sexes may portend greater cardiovascular risk in females compared to males. Until menopause, females are largely protected from atherosclerotic coronary disease, which tends to present clinically a decade later in females compared to males (46). Tobacco cigarette smoking eliminates this female advantage and, in fact, is a stronger risk factor in females; females who smoke 20 cigarettes per day have a 6-fold increased incidence of myocardial infarction, compared to a 3-fold increased incidence in males (46). Thus, our findings that combusted cannabis has similar proatherogenic associations in females and males may herald earlier onset of cardiovascular disease in females, and, like smoking, may erase the protective effects of sex hormones.

Since levels of COS had a strong positive association with foam cell formation in people who used combusted cannabis, and since CB-1 receptor stimulation is known to increase COS production, it is logical to assume that this increase in COS is mediated through CB-1 receptor activation. However, CB-1 receptor-independent mechanisms for COS production have also been identified and should be considered (39). For example, cannabinoids have been found to stimulate TRPV1 receptors (39, 47). TRPV1 stimulation triggers intracellular calcium signaling pathways coupled to ROS generation in mitochondria (48, 49). Additional studies examining specific CB-dependent and CB-independent pathways of COS production are warranted.

The strengths of our study include the inclusion of a healthy, young, and well-characterized population, as well as the use of our novel, mechanistic model of atherogenesis. We also acknowledge certain limitations. Cannabis use and nECIG use frequencies were self-reported, which are always susceptible to recall bias and inaccuracies. To address this, objective markers of substance use were included, such as plasma cotinine levels, scores on the CUDIT-R survey, and urine testing for THC. However, self-reporting remains a common limitation in human studies. Our focus was on individuals who used combusted cannabis at least weekly for the past year, although other forms of cannabis use were permitted. The report by Mohammadi and colleagues included a group who exclusively ingested edible cannabis and observed a significantly reduced FMD response(40). While most cannabis users reportedly use more than one form (50), future research comparing individuals who exclusively use a single form—such as vaping, edibles, or others—would be valuable. Lastly, the toxins and constituents found in smoke generated from cannabis and tobacco largely overlap, except for the cannabinoid and nicotine constituents. Future acute exposure studies involving research cannabis cigarettes with 0% delta-9-tetrahydrocannabinol (THC), followed by known levels of THC, may help clarify the roles of THC versus non-THC constituents in cannabis smoke as mediators of proatherogenic effects. Many of these limitations present opportunities for future investigation.

In summary, chronic cannabis smoking is associated with pro-atherogenic changes in otherwise healthy young people, and the addition of nECIGs further increases these changes. These findings add to the accumulating evidence that chronic combusted cannabis use with or without nECIGs is not harmless.

## Data Availability

The data that support this study's findings are available from the corresponding author upon reasonable request.

